# Developmental and operationalisation influences of Malawi’s Health Sector Strategic Plan III 2023-2030: A qualitative study of the context, processes, content and actors

**DOI:** 10.1101/2025.05.17.25327769

**Authors:** Emilia Connolly, Francis Zhuwao, Anat Rosenthal, Louise Tina Day, Luckson Dullie, Dominic Nkhoma, Tanya Marchant, Gerald Manthalu

## Abstract

**Introduction:** Malawi’s Ministry of Health (MOH) launched the Health Sector Strategic Plan III (HSSP III) 2023–2030, which features catalytic efficiency and accountability reforms under the “One Plan, One Budget, One Report” framework. This policy analysis examined the context, historical influences, processes, and stakeholders shaping the development and early implementation of the HSSP III.

**Methods:** Between June and December 2023, we conducted a document review guideline (n=21) and in-depth semi-structured interviews with health stakeholders (n=12). We initially analysed the data using inductive content analysis, employing the HSSP III development process, followed by a deductive approach that utilised the Walt and Gilson policy triangle, complemented by other conceptual frameworks, with triangulation and consensus validation.

**Results:** Historically, economic and decision-making inequities in Malawi’s health sector have led to centralised processes, vertical funding, and low staff motivation. In response, HSSP III was aligned with current movements for universal health coverage and donor aid coordination. The plan’s development promoted capacity building, country ownership, and accountability through an inclusive and participatory approach, as well as a performance management-oriented framework. However, incomplete operational plans and misaligned decision-making systems have impeded early implementation.

**Discussion/Conclusion:** The Malawi HSSP III responded to the fragmented health sector through a government-led, sector-aligned approach. Its development leveraged long-term leadership and a consultative learning environment to foster capacity and stakeholder ownership. However, the transformative development process alone was not sufficient. Operationalisation and long-term reform require increased government accountability. These findings provide valuable insights for policy development, implementation, and long-term transformational change in Malawi and similar settings.

## Introduction

Sustainable Development Goal (SDG) target 3.8 is to ‘Achieve universal health coverage (UHC), including financial risk protection, access to quality essential healthcare services, and safe, effective, quality, and affordable essential medicines and vaccines for all by 2030.’^1^ As countries strive to achieve UHC, they must prioritize interventions to strengthen their health systems, expand access to health services, and deliver high-quality services.^2^

Health sector countries develop strategic plans (HSSPs) to address this prioritization challenge.^3^ HSSPs are high-level national documents that can reflect 1) shared vision, values, and goals, 2) need and demand for health services, 3) resource prioritisation and allocation toward UHC objectives, and 4) implementation arrangements.^2^ For HSSPs to be effective, they require strong governance to hold stakeholders accountable for achieving and sustaining anticipated system changes and reforms. However, HSSPs’ implementation can be a leadership and technical challenge, especially in the context of limited resources and fragmented health financing arrangements with substantial donor dependence.^3–5^

Countries in sub-Saharan Africa have generated a growing body of literature on health policy development and analysis. Analysis of HSSPs reveals that they often share design elements and face similar planning and implementation challenges, including a need for more contextually relevant evidence and poorly coordinated efforts among governments and stakeholders.^3–7^ HSSP policy reviews have described HSSP development as a complex interplay between historical and current contexts involving multiple actors, including the government, donors, professional organisations, and other health stakeholders. This results in difficulty in executing aspirational policy values and content, as well as challenges in demonstrating HSSP results. Challenges include ineffective planning processes, reliance on donor funding and external technical capacity, and limited use of evidence-based research, policy, and absorptive capacity.^3–5^

The Malawi Ministry of Health (MOH) introduced national health strategies over 40 years ago, with the most recent HSSP II running from 2016 to 2021. Health sector funding, influenced by the configuration of international donor funding and mistrust in public financial management systems,^5, 8^ is mainly in disease-specific vertical programs, such as HIV and malaria, leading to resource duplication.^9^ In response, the Malawi Health Sector Strategic Plan III (HSSP III), launched in 2023, adopted a “One Plan, One Budget, One Report” approach to advance harmonisation and alignment. The plan was developed using multisectoral evidence and consultations with over 200 stakeholders from the MOH and the broader health sector. It aims to unify health sector planning, budgeting, and reporting, with a focus on performance and long-term government budget support.^9^ In the early stages of implementing HSSP III, there is a need to understand the current critical influences, interactions, and interests to optimise implementation efficiency and effectiveness.^4, 5^ This health policy analysis aimed to elucidate the context and actor influences that affect the agenda-setting, content, and processes of the Malawi HSSP III development, and to propose measures to enhance effective implementation.

## Methods

### Study setting

Malawi is a landlocked country in southeastern Africa with an estimated population of 20.3 million, of which 61% are under the age of 25. The Government spends approximately 8-10% of its budget on health. It substantially depends on international aid funding: 58.6% of the total health expenditures come from donors, and over 80% of donor funding is tied to disease-based projects.^9^

The public sector is the principal provider of health services in a three-tiered health system, comprising primary care (including community health), secondary care in the 29 districts, and five tertiary hospitals. Essential health services are free at the point of access at public facilities. Some services, particularly for pregnant women and children, are provided free of charge through service-level agreements with the Christian Health Association of Malawi (CHAM) and the Islamic Health Association of Malawi (IHAM), due to a lack of public facilities. However, stockouts of medicines and commodities, as well as poor health service infrastructure, challenge care delivery. Despite these limitations, Malawi has registered notable health gains in the last 20 years, especially in maternal and neonatal health, malaria mortality, and close achievement of HIV epidemic control.^9^

### Conceptual Frameworks

We conceptualised our study using two frameworks, complemented by power^10^ and political analyses,^11^ as well as social construction frameworks that have been used previously in similar contexts.^12^ First, we used Walt and Gilson’s policy triangle^13^ to emphasise the interplay between the content of the policy and the strong influence of historical and current context, actors, and processes in its development and implementation, aligning with the study’s aim (Figure 1). Second, the 5C approach for policy implementation, as proposed by Cloete and de Coning, supplemented policy development analysis. It links closely to the policy triangle components, including the 1) content of the policy to express how the policy aims to solve problems, 2) context of the institution(s), 3) commitment of implementers, 4) capacity of the administration and implementers and lastly 5) coalition interests.^14^

**Figure 1.**
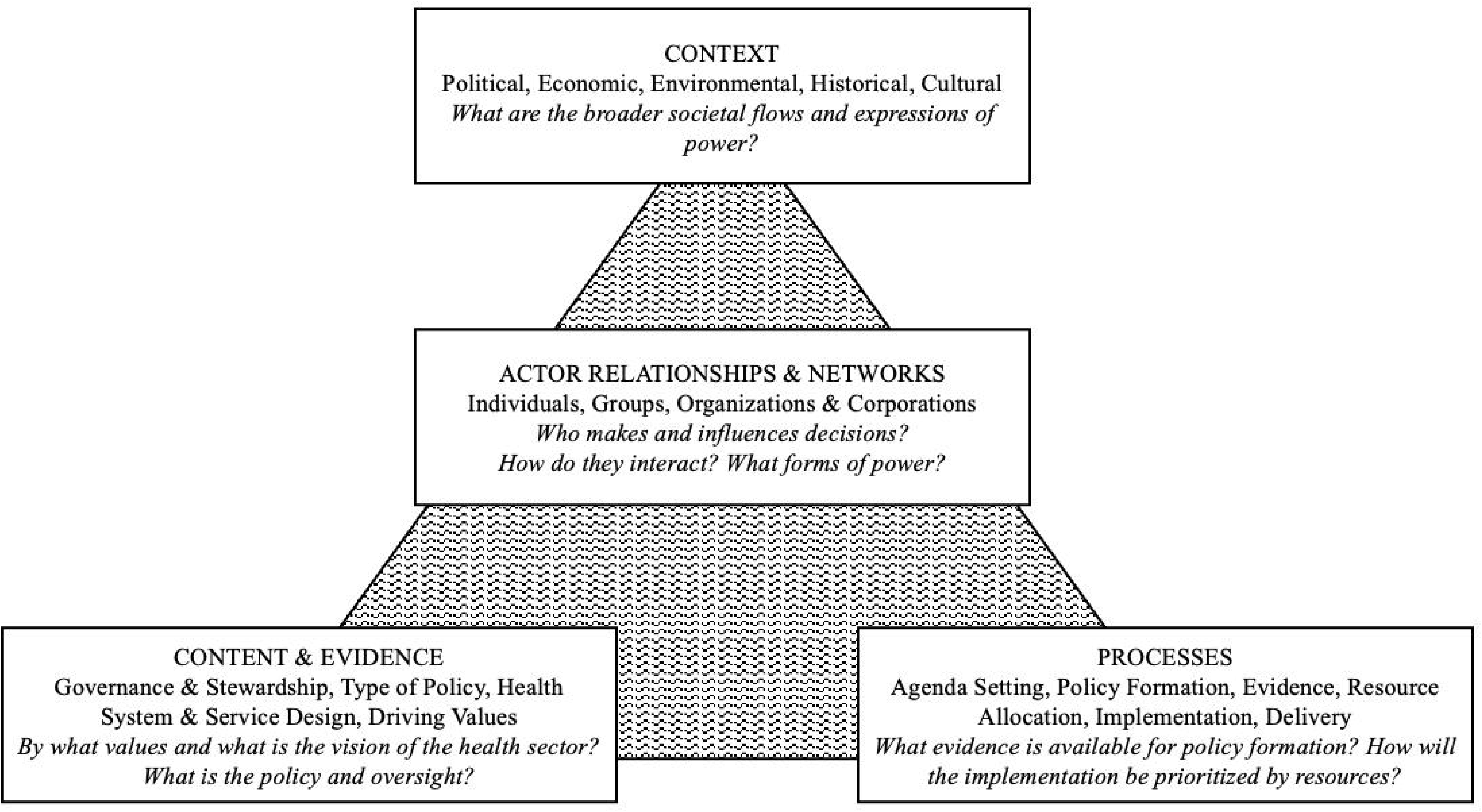
Adapted Walt and Gilson Health Policy Triangle^10, 13^

### Study Design

We employed a cross-sectional, observational, and qualitative approach for this health policy analysis, allowing for triangulation between different sources of evidence. First, from June to September 2023, we did a literature and document review of publicly available recent documents, strategies, and policies from the government of Malawi, donors, and other health policy stakeholders. Second, from August to October 2023, we conducted qualitative in-depth interviews (IDIs) with key informants to describe the contextual and critical influences, organisational factors, and external relationships that facilitated or challenged the development of the HSSP III. An initial stakeholder mapping was completed through a desk review, and participants provided valuable stakeholder insights and input. The study was conducted during the first 10 months of HSSP III implementation.

### Subjects, recruitment, and sample size

#### Desk Review

Documents for desk review were identified by snowballing from existing MOH documents and other organizational resources, with recommendations from MOH and the Health Donor Group (HDG) of Malawi. Criteria for inclusion were current use for policy implementation, evaluation, or data use and publicly available.

#### Qualitative interviews

The IDI methodology was selected to focus on individual stakeholder perceptions and views. The target population was health stakeholders involved with HSSP III formulation (2021 to 2023), including Malawi MOH policymakers, donors, development partners, academics, implementors, and civil society. The inclusion criteria were that the actor had participated in the agenda-setting or development of the HSSP III, was at least 18 years old, spoke English or Chichewa fluently, and had consented to participation. A purposeful and chain referral sampling technique was employed, utilising an HSSP III stakeholder list and soliciting referrals from interviewees. The sample size was based on reaching theoretical (in sampling) and data (in data collection) saturation in qualitative IDI, with saturation met at 12 interviews.^15^

### Positionality and Reflexivity

The study team positionality within this policy analysis varied, including some investigators with specific roles within the Malawi healthcare sector and the HSSP III development. The team consisted of those living in Malawi: Malawians with leadership roles within the MOH, academic institutions, and an international non-governmental organisation and a non-Malawian who has worked in government and research for more than eight years in Malawi and living outside Malawi: non-Malawian academic educators and researchers. Although the study team was gender-balanced, it primarily comprised more senior leadership from their respective organisations.

Acknowledging that this positionality could influence and bias data collection, the choice of conceptual frameworks, and analysis, the study team repeatedly reflected with each other, experts within MOH, and health stakeholders in Malawi.^16^ For data collection, IDI participants were informed before the interview about the strengths and weaknesses of the interviewer’s and study team’s positionality to minimise the social acceptability of interview responses. In data analysis, the study employed triangulation of data with an external review of results to minimise confirmation and anchoring biases.

### Data Collection and Management

#### Desk Review

We reviewed 21 Malawi health sector documents, including donor and stakeholder cooperation strategies, government and professional strategies, operational plans, policies, and health sector reports. One researcher (EC) initially read all documents, taking notes on the author’s category and listing stakeholders, content, and wording, phrasing them according to the conceptual framework.

#### Qualitative Interviews

Data were collected using an IDI interview guide, developed with open-ended, emotionally neutral questions guided by the conceptual framework tenants and a desk review (Supplemental Table 1). The interview guide included questions on the participant’s involvement in the HSSP III, their perception of actor roles and context, stakeholder mapping, facilitators, challenges and turning points of the HSSP III process, and expectations and perceived challenges for implementing the HSSP III. It was developed in English and translated into the local language, Chichewa, for participants who desired to use it. The interview guide was pre-tested with two national-level health sector policy actors. The pretesting results were not incorporated into the findings, but they assisted in refining the questions for clarity and incorporating additional probes.

Ten interviews were conducted in person in a private setting, and two were conducted on a virtual platform by EC. The interviews took approximately 45-70 minutes to complete. All interviews were conducted in English, as requested by the respondents, and were audio recorded. Reflexive memos were completed following each interview. Interviews were initially deidentified using a unique study code for each participant and transcribed using *Trint*.^17^ Interviews were transcribed verbatim with a naturalised approach, incorporating fillers, cut-offs, expressions of emotion, background noises, and interruptions. Transcripts were stored on a password-protected virtual platform in accordance with general data protection regulations. Personal identifiers were replaced with non-specific descriptions. Periodic debriefs of transcriptions were conducted with the study team throughout data collection to ensure rigour. Data were managed on *Nvivo 12.* All data were stored on a password-protected, encrypted computer hard drive with access only for EC.

### Data Analysis

Document review, stakeholder mapping, and IDI data were coded in *Nvivo 12.* The preliminary codebook was initially developed by EC using an open inductive HSSP III development timeline approach, allowing natural themes to emerge during the preliminary coding round. Following the preliminary coding round, two independent investigators (EC and AR) revised the codebook based on feedback from the initial analysis and simultaneously coded sample interview transcripts to resolve discrepancies in coding. All authors agreed on the updated codebook through a joint consensus, using 17 codes to code all documents and transcripts (Supplemental Figure 1). EC completed coding on all transcripts using the validated codebook, with cross-coding for 20% of interviews completed by AR and GM. Excerpts and quotes were exported into a Word document for easy immersion and/or familiarisation with the data through repeated and active reading, as well as connections between themes. Finally, two researchers (EC and AR) collaborated to identify relationships between codes, identify frequent codes, and generate themes across transcripts and documents, incorporating input and review from other authors. We employed an inductive approach to identify emerging thematic categories that align with the Walt and Gilson policy triangle.^13^ The 5C’s of policy implementation by Cloete and de Coning^14^ complemented deductive analysis for operationalising the HSSP III.

Following coding and thematic analysis, we triangulated the analysis between the document review, stakeholder mapping (Supplemental Figure 2), literature review, and IDIs. Selected study team members (EC, GM, FZ, and DN) reviewed the triangulated analysis with respondents and selected stakeholders in a one-on-one or pairs format to provide expert feedback and insight into the findings and recommend additional and adjusted themes. This process created consensus and validation for the final analysis and results.

### Ethics

This study was approved by the National Health Science Research Committee (NHSRC) in Malawi, protocol number 23/03/4019, and the London School of Hygiene & Tropical Medicine (LSTHM) Ethics Committee in the United Kingdom, protocol number 29007. All respondents were above 18, received a participant information sheet, and signed written consent for participation and audio recording.

## Results

We reviewed 21 Malawi health sector documents and interviewed 12 health stakeholders and actors involved in the formulation of HSSP III. The documents included five (5) donor and stakeholder cooperation strategies and reports, twelve (12) government strategies, operational plans, guidelines, and policies, three (3) health surveys and reports, and one (1) professional organisation code of conduct. The documents are presented in Supplemental Table 2 with a content summary for each. The interview sample consisted of 7 (58%) males and 5 (42%) females, with 6 (50%) affiliated with the Ministry of Health and the remaining 6 (50%) representing other health sector stakeholders (Supplemental Table 3). Of the interviews, 5 (42%) were trained in clinical medicine or nursing, 6 (50%) were trained in economics or policy, and 1 (8%) were trained in a related field, with half (50%) holding a doctorate. Finally, 4 (33%) of the interviewees had worked in the health sector for less than 10 years, 6 (50%) had worked for 10-20 years, and 2 (17%) had more than 20 years of experience in the health sector.

Four main themes emerged from the coding and analysis of the document review and interviews examining the development and initial implementation of the HSSP III, including 1) the historical context, actors, and power in policy-making; 2) the current policy environment focusing on actors and practices; 3) the development process of HSSP III with changes in process and content; and 4) challenges and enablers of operationalising the HSSP III (Supplemental Figure 3). Within these categories, we analysed the data to explore power and governance imbalance between actors within Malawi’s perception as a “donor darling,” the current health sector environment of fragmentation, inconsistent decision-making, and lack of incentives for government capacity, the HSSP III development process focused on learning and accountability for mindset and system change and enablers and challenges of HSSP III implementation. The quotes in the results section represent the overall data set and highlight major themes and trends from the document review and interviews.

### 1. Historical context and actors in policy making in Malawi

The historical context, power structures, and stakeholder perceptions have shaped policy development and implementation, which are influenced by government debt and aid dependency, as well as bureaucratic inefficiencies. At the same time, Malawi is often seen as a “donor darling” for external stakeholders.

#### 1.1. Power and governance imbalance between actors

Despite over 50 years of a Malawian independent government, there remain competing power dynamics, strategic decisions, and policies with external donors and actors due partly to past colonialisation.^5^ One participant shared:

> *“I think one of the things that [is] very prevalent in Malawi is in other countries as well. This bureaucracy has a history rooted in colonial times. It comes through [in all interactions], and it just rolls on. (P3, stakeholder)*

The power dynamics between the Malawi government and external actors, coupled with a dependence on funding, have influenced decision-making in the policy environment. One participant recognised:

> *We take a backseat role and let them lead. Despite that, they tell you that this is the person you need to be. I don’t know if that’s the dynamics that comes with all the power plays [because they have] the resources and expertise…it’s something we can’t do without for the time being. (P6, MOH)*

#### 1.2. Donor Darling

Within the historical context, Malawi is known for its peaceful and welcoming culture, which has significant resource needs but is open to diverse ideas. These attributes can result in Malawi being sought out by donors, researchers, and implementing partners. One participant stated:

> *Malawi is kind of a donor darling. Malawi is one of the nations favored for innovations. It has always been in the forefront. (P2, stakeholder)*

Another participant commented on the comparatively uncomplicated, primarily public, free health system:

> *In Malawi, some of the systems have already been designed. The state health care is meant to be free. It is easy to conceptualize what should be happening ideally and the various health system levels within the systems. (P4, stakeholder)*

### 2. The current policy environment in Malawi

In Malawi, health sector funding is primarily channelled through program-based initiatives. The existing financing and power imbalance between the Ministry of Health (MOH) and donors, as well as between national and subnational MOH, hinders decentralisation efforts. It reduces civil service incentives, like competitive salaries and autonomous decision-making.^9^ As a result, there is weak ownership of MOH policies and inconsistent decision-making, which leads to fragmentation and resource duplication.

#### 2.1. Fragmentation of the Health Sector

Participants highlighted that vertically funded programs, often aligned with donor priorities, have resulted in multiple meetings and implementation efforts without effective coordination between activities. In line with donor priorities and funding, participants noted that the MOH departments often lack alignment with each other. One participant noted:

> *We talk about donors, but it is the same thing within the ministry. Sometimes, the right-hand does not know what the left-hand is doing…So, things seem to still happen in silos. Better coordination is needed both within and outside the ministry. (P3, stakeholder)*

The lack of coordination between disease-based departments results in siloed activities and duplication of efforts on the ground. One participant stated:

> *Fragmentation has been challenging over the past decade. In the past few years, we have seen how disease-specific interventions tend to be driven by certain agencies and certain donors and how that impacts the work on the ground. That is duplication of effort, that is fragmentation. (P2, stakeholder)*

Finally, participants observed that fragmentation and centralised decision-making often lead to a breakdown of the actual policy execution. One participant shared:

> *[Successful health care interventions] require reallocation of power to the districts… [We need] people who buy into decentralised decision-making and autonomy and are willing to invest and adjust their funding, planning, budgeting, and implementation. (P7, stakeholder)*

#### 2.2. Lack of enforcement and inconsistent decision-making

Participants identified weak enforcement by the MOH and inconsistent decision-making as barriers to policy implementation, often due to funding constraints. External influences, such as consultants and donors, frequently impact decisions, underscoring the need for greater MOH ownership. One participant shared:

> *I think the HSSP III development was run by the government. However, the [HSSP III prioritszation] was influenced a bit by people who had money. If you want to have donor buy-in, you can’t say no to the donor, right? (P7, stakeholder)*

This type of donor-influenced environment may lead to less MOH process ownership, especially when there can be external, often donor-funded external consultants leading processes. One participant remarked on an experience with an external consultant in a policy development process:

> *I thought [the consultant] was trying to defend whatever [he] produced. He was not really hearing what other people were talking about. I think when he went back, he wrote what he wanted rather than be informed by our experience because he seemed to think he was the subject expert in the field. (P6, MOH)*

Interviewees believed that if public servants had access to increased government resources, there would be greater enforcement and accountability by the government. One participant suggested:

> *There needs to be more resource mobilisation for the government. Once they increase the level of funding, the accountability will be stronger from the government offices. (P3, stakeholder)*

Participants noted that the overwhelming number of donor meetings, fragmented initiatives, and external power dynamics have led to inconsistent decision-making. One participant highlighted that MOH decisions are often made ad hoc rather than being guided by established national-level policies:

> *There are two things in the Ministry - policy decisions and the actual policy. I think much more is done through decision-making that does not reference any policy document. (P11, MOH)*

#### 2.3. Lack of incentives for government capacity

Participants noted that poor or misaligned incentives, particularly related to unequal power and funding within national and sub-national Ministries of Health, as well as between the MOH and donors, could lead to a limited capacity among government public servants, often characterised by inadequate numbers and competence. This environment can hinder the ownership and implementation of MOH policies at all levels. Participants emphasised the necessity of decentralised capacity building, especially at the district level, to enhance policy execution:

> *To make central-level ideas practical at the district level…I think a lot of capacity building is needed at the district level to bring them the same kind of vision and strategy thinking that might be originating at the national level. (P2, stakeholder)*

Given the historical and current power imbalance between donors and the Ministry of Health (MOH), alongside a heavily bureaucratic system and centralised decision-making, participants noted that the MOH system may be unwelcoming to new ideas and may discourage civil servants from sharing them. One participant shared:

> *The [MOH] has left me stagnated with little innovation or ability to change the situation of the health care system. People are adamant about how things are. I became a problem because I wanted to do things differently. (P6, MOH)*

The current policy-making environment in Malawi culminates in the feeling of being “stuck in the middle” (P4, stakeholder). This term describes how health stakeholders operate within a democratically elected government, often lacking accountability and follow-through in implementing and evaluating policies. Several HDG documents illustrate this sentiment, for example:

> *Malawi’s development challenges are interwoven and inseparable. Like a tangled mass of interlocking string, pulling on one thread alone may help loosen the problem, but is unlikely to unravel it fully. (USAID National Cooperation Strategy 2020-2025, page 1)*

### 3. Development process of HSSP III

The HSSP III development process focused on individual actors and sector-wide mindset changes to catalyse a health system strengthening approach from the current, verticalised, and siloed system. This was operationalised through an enhanced policy and framework, creating a learning environment with collective ownership and responsibility.

#### 3.1. Actor mindset and systems change

The HSSP III called for a change in mindset to systems change by drawing on historical health system policies within Malawi, national and global recognition of a health systems approach, such as the Futures of Global Health Initiative (FGHI)^18^, and utilising evidence for development. One participant reflected:

> *The [HSSP III] was so well thought through and aligned with so many things. It’s easy to tell people we are doing this because everybody else also thinks the same things. So many processes validate it, such as the FGHI process, which validates the thinking going on there. (P11, MOH)*

Interviewees saw the need to shift from a personal or departmental perspective to a holistic health sector and patient-care benefit mentality. One interviewee commented:

> *I saw that people were more interested in how [activities] benefit them than how they benefit everyone else in the long run. That’s an attitude thing, which is very difficult to tackle. (P6, MOH)*

#### 3.2. Linkage of the HSSP III framework to system reform and performance

Interviewees viewed the HSSP III framework as an improvement over previous HSSPs because it emphasises reform and enhances accountability. Participants discussed the HSSP III reforms related to performance management, expressing hopes to shift their mindset towards a health system-strengthening approach. One participant explained:

> *Using the WHO building blocks, you can [identify challenges] and [solve by] the reforms. The approach was impressed by the need to arrive at an M&E framework, which presents an opportunity to link a performance management framework that you can link to the reforms. (P8, stakeholder)*

With this framework, there was an appreciation for how it shifted the roles and mindset of MOH departments through a health system-strengthening approach. One participant remarked:

> *There was a pivotal change in how the Ministry was thinking about health systems and service delivery in an integrated manner as part of that “One Plan, One Budget, One Report” framework. It was like more alignment integration. So, everything was supposed to be different from what it was and how business as usual has been. (P7, stakeholder)*

#### 3.3. Creating a learning environment

Participants recognised that the HSSP III consultative development intentionally fostered an open and adaptive learning environment for everyone involved. Stakeholders were able to see their contributions reflected in the HSSP III within a space where ideas were challenged, debated, and refined. One participant noted:

> *[The HSSP III] encouraged the culture of people to see new knowledge. Part of learning and un-learning, it’s quite important. (P3, stakeholder)*

This learning environment was enhanced by intentionally including diverse stakeholders, such as civil society, policymakers, and district-level implementers. This participant noted:

> *In the previous strategic plans, you know the engagement was marginal. For the HSSP III, engagement was pushed deliberately. The implementers were supported to build their vision. The sessions ran with two hundred people and built an army of [HSSP III] champions. (P8, stakeholder)*

The HSSP III development process fostered a learning culture, amplifying the voices of implementers and focusing on strengthening the health system. Participants highlighted global health system approaches that influenced HSSP III, with a persistent need for mindset shifts to improve care delivery.

### 4. Operationalising the HSSP III

The HSSP III was launched in January 2023 and became operational immediately. Interviewees observed enablers and challenges to operationalising the “One Plan, One Budget, One Report” approach during the first six to ten months of implementation.

#### 4.1. Enablers of HSSP III Implementation

The enablers of HSSP III execution align with the 5Cs of policy implementation, emphasising a supportive global and national context, aligned interests, and the connection of reforms to performance management. Crucial to its success was the leadership of seasoned MOH policymakers and effective collaborations that united stakeholders. One participant noted:

> *There was discontent with the status quo. The longevity of leadership is also a good factor. We have waited for this moment to do this - the right time and the right platform have come to do this. (P11, MOH)*

#### 4.2. Challenges of HSSP III Implementation

Participants identified ongoing challenges, including the necessity for a committed operationalisation plan for HSSP III that specifies process changes and holds accountability while adjusting resource governance. One interviewee commented:

> *The implementation modalities in the document are not very clear. That leaves room for business as usual. This nice plan may not be delivered effectively because it hasn’t been clear. And [that’s] a recipe for disaster. (P5, stakeholder)*

Participants noted that, despite the HSSP III implementation enablers, there are still remaining challenges, including the need for disseminated operational plans to enhance input efficiency and effectiveness.

## Discussion

This policy analysis highlighted structural inequalities, fragmented efforts, and limited incentives for coordination in Malawi, which challenged policy development, implementation, and evaluation. However, it also highlighted a growing momentum for a health system-strengthening approach to align funding and policies better nationwide. The development of HSSP III fostered a shift toward accountability and performance-focused learning, planning, and implementation in the health sector. Nevertheless, sustaining the coordination gains from HSSP III will require persistent and incremental efforts to address these complex policy and structural issues.

### The complexity of policy development and operationalisation

The Walt and Gilson^13^ policy triangle, initially focused on policy content, actors, processes, and context, has evolved through scholars such as Topp *et al.*^10^ to highlight power dynamics and political networks in policymaking. Participants in HSSP III noted that actors and organisations, including donor and recipient governments, frequently have conflicting interests that influence the policy agenda, process, and content. These competing interests and the context and prior policy development and implementation processes impacted HSSP III’s development and early execution. Changing the visualisation from a triangle to a Venn diagram may better illustrate the complex overlaps between actors, context, and content, highlighting the interdependent and often messy nature of policy development and implementation (Figure 2).

**Figure 2.**
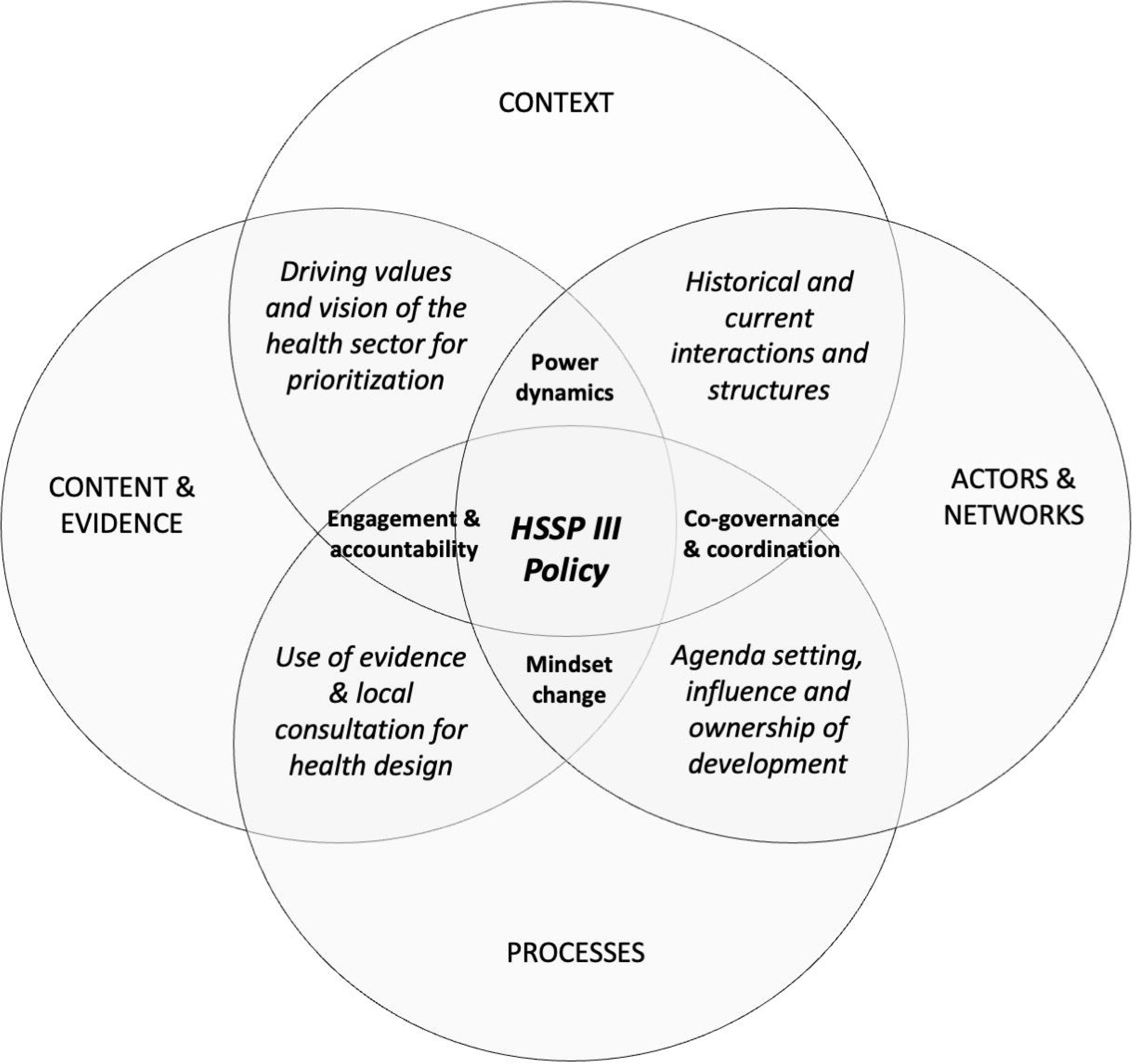
Venn diagram demonstrating the complexity of the Malawi HSSP III development through the policy triangle framework

### Context and key influences of the “One Plan, One Budget, One Report” approach

Our findings highlight how the legacy of colonialism and ongoing aid dependency have shaped Malawi’s current governance and policy environment, contributing to a centralised, bureaucratic system. This has led to inconsistent financial management, donor funding flows, and a lack of trust among health stakeholders despite consistent past MOH policies and funding utilisation.^8, 19^ Most donor funding is directed towards specific disease areas, such as HIV, as reported in several similar contexts.^7^ This results in fragmented planning and reporting, as evident in Malawi’s over 50 strategic plans.^9^

Despite this, participants noted positive global, national, and sub-national efforts to coordinate better government and donor funding, planning, and implementation, leading to increased government ownership and accountability. Several global frameworks, such as the Paris Agreement^20^ and the Lusaka Agenda,^18^ have influenced Malawi’s initiative to align planning and implementation in the current HSSP III. Several African countries, including Ethiopia^21^, Burkina Faso, and Rwanda^22^, have previously adopted a “One Plan, One Budget, One Report” approach to help unify their strategic vision and shift accountability to the government. Similar pooled donor funding and sub-national direct support have been introduced in Malawi, although managed by separate management and implementing teams to align with donor funding streams. For example, there is a Health Services Joint Fund (HSJF) with pooled donor funds earmarked for government priorities but managed by a separate contracted oversight team. All spending proposals must be vetted and approved by the donors.^23^

However, the results indicate that to fully implement the HSSP III, existing governance, accountability, and decentralisation must be addressed. A single aligned framework or short-term changes cannot solve Malawi’s complex social, political, and power dynamics.^24^ The HSSP III’s consultative, reform-based approach represents a step toward change. However, implementing this plan encounters significant challenges due to entrenched donor interests, centralised power structures, resource shortages, and systemic barriers, as illustrated in Ethiopia.^21^ Incomplete operational plans and a lack of transparent enforcement hinder efforts to redefine government and health sector processes. Moving forward, Malawi must adopt a consistent, iterative approach to tackle these challenges and achieve a government-led, coordinated health sector.

### Organisational and relationship factors in HSSP III process development

The Grindle and Thomas framework^11^ and social construction theory^12^ illustrate that policymakers, donors, and implementers operate in a complex environment of values, interpretations, and power. While these actors believe they are guided by evidence, they often interpret information to fit their preconceived notions. This can lead to contradictory dynamics in policymaking. For example, Malawi, known as a “donor darling” for its funding for progressive initiatives like the rollout of dolutegravir – the first integrase inhibitor for HIV treatment publicly available in Malawi - faces issues such as stakeholder fragmentation and power imbalances.^25^ Although 35% of funding for HIV comes with specific donor mandates,^26^ the relationships between governments and stakeholders indicate that policymaking is not simply a “zero-sum game” (where one side has resources and the other does not).^27^ Our findings and existing literature highlight the need for policymaking to focus on distributive, contextualised government ownership for sustainable development.^4, 6, 28^

The HSSP III development process employed a consultative and adaptive approach led by the Ministry of Health (MOH). This strategy aims to shift from reliance on external consultants to enhancing local leadership, fostering ownership, and strengthening incentives across all levels of the health system.^24, 29^ Interviewees noted that external “expert” consultants in Malawi often lead to a lack of ownership and a sense of being cornered, a phenomenon referred to as “*consultocracy.*”^29^

Furthermore, centralised control within the MOH restricts district-level strategic thinking, priority setting, and accountability—issues also evident in Uganda, Kenya, and Ghana.^19, 28, 30^

To address this, the HSSP III seeks to reduce dependence on consultants and boost sub-national participation. This shift aims to balance power, facilitate knowledge transfer, and empower civil servants to solve problems effectively, thereby ensuring the successful implementation of long-term policies.^24, 29^ However, while this approach was adopted in developing HSSP III, its execution has lacked clear operational plans and commitment. Moving forward, the emphasis should be on collaborative policy implementation and fiscal decision-making between the central MOH and local communities rather than a top-down model from the central MOH and donors.^24^

### Strengths and Limitations

This study is the first review of HSSP III, focusing on its ongoing operationalisation and enhancing alignment within the health sector. Although conducted in one country and with one policy, the findings align with existing literature on other HSSPs and policies in Malawi^5, 8, 19^ and the region,^3, 4, 6, 7^ highlighting similar actors, networks, contexts, and processes within the policy triangle.

A limitation of the study is its cross-sectional design, which provided time-bound data. Data collection began seven months after the launch of HSSP III, limiting participants’ recall of earlier events. Additionally, gathering data within a year of HSSP III implementation restricted the monitoring of ongoing implementation and changes in perspectives. Finally, the individual interviews within a small policy space may have introduced biases such as social acceptability and confirmation bias. However, these were mitigated by triangulating interviews with desk and stakeholder reviews.^15^

## Conclusion

The Malawi HSSP III (2023-2030) addresses a complex and fragmented health sector through a government-led, sector-aligned “One Plan, One Budget, One Report” approach. Despite structural inequities, HSSP III leverages global promotion to align UHC and MOH leadership, creating a performance-based framework that encourages a decentralised, growth-oriented learning environment to shift stakeholder attitudes. Persistent power structures, verticalisation, and limited accountability planning challenged its development and operationalisation. Success requires embedding individual and departmental accountability within the government, along with a realigned reporting framework for performance management and long-term change. These findings can inform Malawi, other countries, and international policy makers on how to enhance sector alignment and quality. Further research should examine long-term sector performance, patient outcomes, policy impacts, and health sector strategic plan operationalisation bottlenecks.

## Abbreviations

AIDS: Acquired Immunodeficiency Syndrome
CHAM: Christian Health Association of Malawi
HDG: Health Donor Group includes World Health Organization (WHO), United Nations Children’s Fund (UNICEF), United Nations for Population Activities (UNFPA), Foreign, Commonwealth, and Development Office of United Kingdom (FCDO), United States Agency for International Development (USAID), Centers for Disease Control (CDC), Kreditanstalt für Wiederaufbau (KfW)/ Deutsche Gesellschaft für Internationale Zusammenarbeit (GIZ), Norway, World Bank/ Global Financing Facility (GFF), Joint United Nations Programme on HIV/AIDS (UNAIDS), and World Food Programme (WFP)
HIV: Human Immunodeficiency Virus
HSSP: Health Sector Strategic Plan
HSJF: Health Sector Joint Fund
IDI: In-depth interview
IHAM: Islamic Health Association of Malawi
KUHeS: Kamuzu University of Health Sciences
LMIC: Low-and middle-income countries
LSTHM: London School of Hygiene & Tropical Medicine
M&E: Monitoring and Evaluation
SDG: Sustainable Development Goals
UHC: Universal Health Coverage

## Author contribution statement

The authors confirm their contribution to the paper as follows: study conception and design: EC, GM, FZ, DN, AR, TM, LTD, LD; data collection: EC; data analysis: EC, GM, AR; interpretation of results: all authors; draft manuscript preparation: EC; critical revision of the article: all authors. All authors reviewed the results and approved the final version of the manuscript.

## Acknowledgements

We thank the Malawi Ministry of Health for sponsoring this policy review and supportive Ministry of Health colleagues, especially within the Department of Planning and Policy Development. Thank you to all the interviewees for participating in this study, as well as to the many academic and implementing partners, donors, and civil society stakeholders who reviewed and refined the study’s outputs.

## Data Availability Statement

The complete datasets generated and analysed during the current study are not publicly available due to participant privacy. However, the aggregated data set can be obtained from the corresponding author upon reasonable request.

## Supplemental Files

*Supplemental Figure 1. Code analysis through the distribution of total reference codes by frequency*

*Supplemental Table 1. Current Malawi Health Sector Documents included in the desk review of the HSSP III policy review (n=21 documents)*

*Supplemental Table 2. Interview Participant Characteristics (n=12 interviews)*

*Supplemental Figure 2. Framework analysis thematic categories*

*Supplemental Table 3. In-depth Interview guiding questions linked to the conceptual framework, participants, and document review*

*Supplemental Figure 3. HSSP III Stakeholder Involvement Mapping*

