## Supplemental Figure 1 for "Developmental and operationalisation influences of Malawi’s Health Sector Strategic Plan III 2023-2030: A qualitative study of the context, processes, content and actors"

*Figure 2. Code analysis through the distribution of total reference codes by frequency*

Reference Code Frequency

| Anaysis Codes | Total Reference codes |
| --- | --- |
| Actor and Organization Interactions | 130 |
| Government and stakeholder capacity | 50 |
| Power and influence in health sector decision making | 148 |
| Fragementation and poor coordination | 10 |
| Health systems strengthening approach shift | 10 |
| Political governance and accountability | 91 |
| Historical and current policy development | 148 |
| Strategy leadership and ownership | 147 |
| Coordination and engagement | 99 |
| Capacity building in health sector decentralisation | 105 |
| Evidence process and use in strategic plans | 72 |
| Effect of fiscal space on health strategy and implementaton | 69 |
| Health system shocks | 5 |
| Lack of inertia and fatigue in health sector | 11 |
| Mindset change for service delivery | 48 |
| Enablers and barriers of HSSP III implementation | 28 |
