## Supplementary figures and images for "Developmental and operationalisation influences of Malawi’s Health Sector Strategic Plan III 2023-2030: A qualitative study of the context, processes, content and actors"

### Supplemental Figure 2

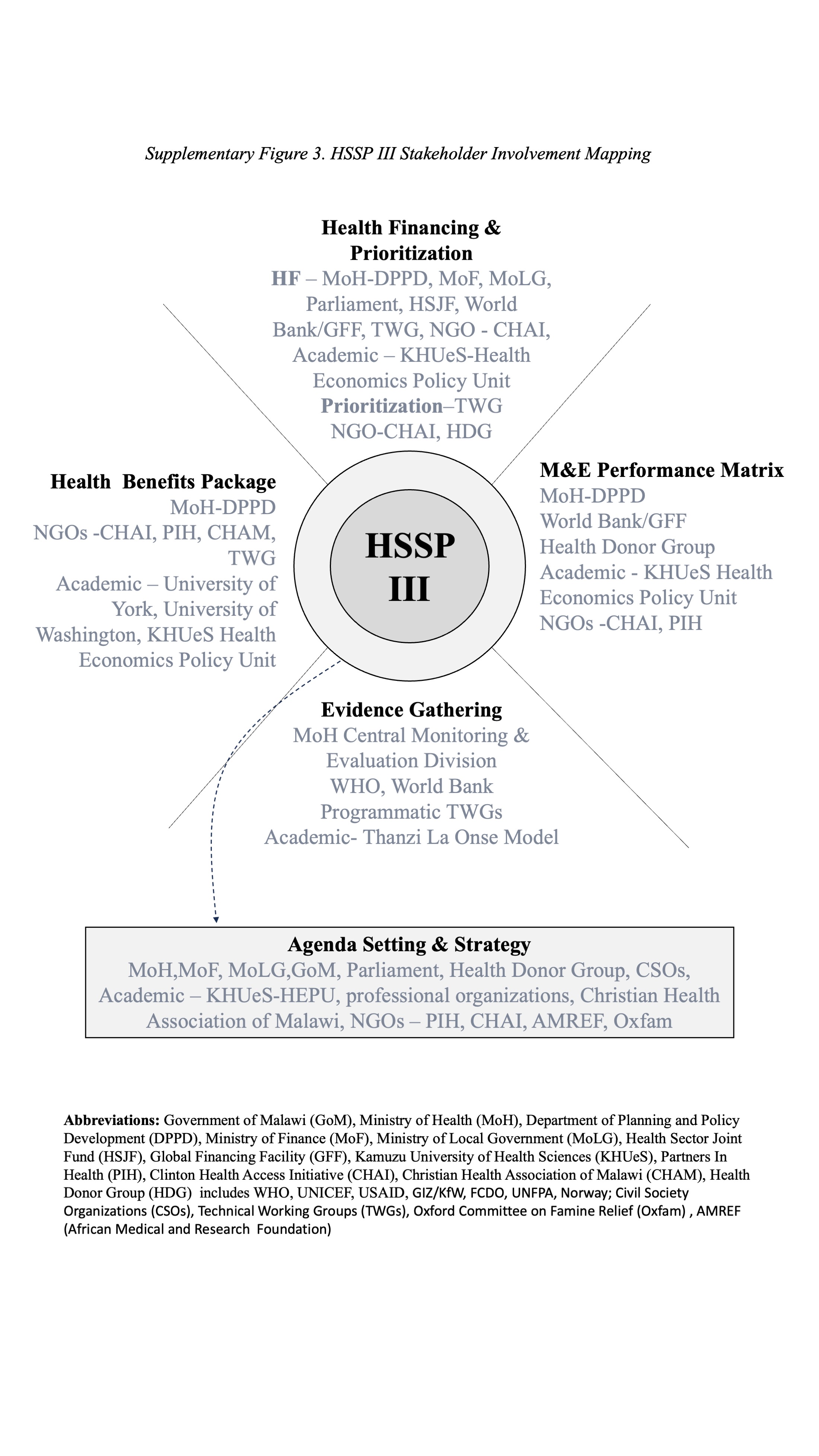
