## Supplemental Figure 3 for "Developmental and operationalisation influences of Malawi’s Health Sector Strategic Plan III 2023-2030: A qualitative study of the context, processes, content and actors"

Figure 3. Framework analysis thematic categories

|  |  |
| --- | --- |
| Historical context, actors and power in policy-making | 23.7404% |
| Current policy environment | 26.3877% |
| HSSP III development process | 47.4808% |
| HSSP III initial opertionalization | 2.39112% |
