## Supplemental Table 1 for "Developmental and operationalisation influences of Malawi’s Health Sector Strategic Plan III 2023-2030: A qualitative study of the context, processes, content and actors"

*Supplemental Table 1. In-depth Interview guiding questions linked to the conceptual framework, participants and document review*

| **Question** | **Conceptual framework linkage^30,31^** | **Potential stakeholders to be interviewed** | **Potential documents for review** |
| --- | --- | --- | --- |
| 1. How do you come to this role? What was your training, journey, and experience in policy processes throughout your professional career? What policies have you been involved with in the past? | Actor relationships and networks  Context | - Central and sub-national Ministry of Health (MOH) - District and Tertiary Care Delivery - Ministry of Health - Academic stakeholders - Donors /Development Partners/ Implementation Partners - Civil society representatives | - Health Sector Strategic Plan III (and past strategies), Malawi Vision 2063 and other governmental strategy documents - Donor and development partner strategic plans and documents/ assessments - MOH Departmental Strategic Plans and Guidelines - Academic policy projects and literature in Malawi - Published and non-published reviews and assessments of HSSP - Technical Working Group and HSSP III Secretariat TOR and documentation - HSSP III consultation reports - Stakeholder analysis - Funding documentation/ resource mapping - Technical capacity building contracts and documents |
| 2. What has been your involvement in development of the HSSP III? How were you involved – certain part of the process or section of the HSSP III? Who invited you to be involved? | Actor relationships and networks  Institutions  Processes  Context |  |  |
| 3. What type of evidence do you regard as most valuable for development of the HSSP III and prioritization? What about others in the Ministry of Health? What about in other organizations? Where do you turn for the information? | Actor relationships and networks  Processes  Content & evidence  Context |  |  |
| 4. Were there any turning points or “Ah-Ha moments” or events that shaped the HSSP III? | Actor relationships and networks  Processes  Content & evidence |  |  |
| 5. Where there moments of difficulty or challenge in development of the HSSP III policy? How were they resolved? | Actor relationships and networks  Processes  Content & evidence  Context |  |  |
| 6. What was the role of donors (or development partners or implementing organisations or academic or regulatory bodies or Ministry of Health departments/ districts/ central hospitals)** in the policy process?    a. Would you help me map stakeholders in the HSSP III process? Which stakeholders had a large role? Which ones perhaps should have had a large role? In what areas of development? | Actor relationships and networks  Processes  Context  Commitment/interest of individuals, institutions and coalitions  Capacity of administration and implementers |  |  |
| 7. Would you do anything differently looking back in the policy process? What would you do different in the future or with iterations of the HSSP III? What are the most important things? | Actor relationships and networks  Processes  Content & evidence  Context  Commitment/interest of individuals, institutions and coalitions  Capacity of administration and implementers |  |  |
| 8. Do you think this HSSP III will work better than past HSSP? How? If yes, why will it work better now? If no, what would make it work better? | Processes  Content & evidence  Context  Commitment/interest of individuals, institutions and coalitions  Capacity of administration and implementers |  |  |
| 9. What do you think health care in Malawi will look like at the end of the HSSP III? Same challenges? What will improve? | Actor relationships and networks  Processes  Content & evidence  Context  Commitment/interest of individuals, institutions and coalitions  Capacity of administration and implementers |  |  |
