## Supplemental Table 2 for "Developmental and operationalisation influences of Malawi’s Health Sector Strategic Plan III 2023-2030: A qualitative study of the context, processes, content and actors"

*Supplemental Table 2. Current Malawi Health Sector Documents included in the desk review of the HSSP III policy review (n=21 documents)*

| **Author Category** | **Title and enacted dates** | **Content Summary** |
| --- | --- | --- |
| Donor and stakeholder | World Bank: Country Partnership Framework for The Republic of Malawi for the Period Fiscal Year (FY):21-25 | *Overall country engagement and strategies of World Bank with Malawi government including the health sector* |
|  | USAID: Country Development Cooperation Strategy (CDCS): April 2020-2025 | *USAID country strategy and goals in Malawi including the health sector* |
|  | Germany Agency for International Cooperation *(*GIZ) in Malawi: Shaping Sustainable Development, 2019. | *Overview of Germany Agency for International Cooperation engagement in Malawi* |
|  | World Health Organization Country Co-operation Strategy 2017-2022 | *Strategic agenda for WHO cooperation in Malawi* |
| Malawi government | Malawi’s Vision: an inclusively wealthy and self-reliant nation (Malawi 2063) 2020 | *Malawi’s vision to be an upper-middle-income country by the year 2063 with goals and strategies for achievement* |
|  | National Health Policy: “Towards Universal Health Coverage” 2018 | *Policy direction, linkages and priority areas with implementation arrangements* |
|  | Health Sector Strategic Plan III 2023-2030: Reforming for Universal Health Coverage | *Overall health sector strategies and reforms for shaping operations and care delivery* |
|  | Emergency and Critical Care Strategy: Framework for Implementing Emergency & Critical Care Services in Malawi 2021-2031 | *Strategies, implementing arrangements and framework for strengthening emergency and critical care services* |
|  | National Tuberculosis (TB) and Leprosy Control Strategic Plan 2021-2025 | *Mission, goals, and strategies to eradication of tuberculosis and leprosy* |
|  | National Health Communication Strategy 2021-2026 | *Strategies and activities for health promotion and prevention activities to address social risk factors* |
|  | Malawi National Sexual and Reproductive Health Rights (SHRH) Strategy 2017-2022 | *Strategies, guiding principles, and activities to implement SRHR* |
|  | National Malaria Strategic Plan 2023-2030 | *Agenda setting and interventions for eradication of malaria* |
|  | National Strategy Plan for HIV and AIDS 2020-2025 | *Strategic planning, resource inputs and interventions for HIV/AIDS epidemic control* |
|  | National Community Health Strategy 2017-2022 | *Outlining the community health framework and system with implementation plan* |
|  | Quality Management Policy 2017 | *Outlining priority areas for quality management with institutional and implementation arrangements* |
|  | Digital Health Strategy 2020-2025 | *Agenda and strategies to strengthen digital health with priorities and implementation activities* |
| Health Sector Surveys | Malawi’s Demographic and Health Survey (2015-16 MDHS) | *Household and individual level data on a broad range of service utilization, mortality, and morbidity* |
|  | Malawi Multiple Indicator Cluster Survey (MICS) 2019-2020 | *Sociodemographic survey of households and individuals on health and health-related practices* |
|  | Malawi Harmonized Health Facility Assessment (HHFA): 2018-2019 Report | *Health facility assessment on service availability, readiness, quality of care, client experience and facility management* |
| Professional Organization Documents | Medical Council of Malawi Code of Ethics and Professional Conduct | *Code of conduct for medical providers on expected duties, responsibilities, relationships, private and advertising partnerships, and emerging issues such as sharing patient and work information on social media* |
