## Supplemental Table 3 for "Developmental and operationalisation influences of Malawi’s Health Sector Strategic Plan III 2023-2030: A qualitative study of the context, processes, content and actors"

*Supplemental Table 3. Interview Participant Characteristics (n=12 interviews)*

| **Characteristic** | **Frequency (n)** | **Proportion (%)** |
| --- | --- | --- |
| ***Gender*** | | |
| Male | 7 | 58% |
| Female | 5 | 42% |
| ***Role affiliation*** | | |
| Ministry of Health | 6 | 50% |
| Other health stakeholder | 6 | 50% |
| ***Level of training*** | | |
| Bachelors degree | 3 | 25% |
| Masters | 3 | 25% |
| Doctorate | 6 | 50% |
| ***Training type*** | | |
| Clinical/nursing | 5 | 42% |
| Economics/policy | 6 | 50% |
| Research | 1 | 8% |
| ***Health sector experience*** | | |
| <10 years | 4 | 33% |
| 10-20 years | 6 | 50% |
| >20 years | 2 | 17% |
